# The role of the H_2_S and OT systems in pre-eclampsia

**DOI:** 10.1101/2025.06.06.25329101

**Authors:** Tamara Merz, Sarah Ecker, Nicole Denoix, Oscar McCook, Stefanie Kranz, Ulrich Wachter, Edit Rottler, Thomas Papadopoulos, Christoph Fusch, Cosima Brucker, Jakob Triebel, Thomas Bertsch, Peter Radermacher, Christiane Waller

**Affiliations:** Institute for Anesthesiological Pathophysiology and Process Engineering, Ulm University Medical Center; Clinic for Anesthesiology and Intensive Care, Ulm University Medical Center; Department of Psychosomatic Medicine and Psychotherapy, Nuremberg General Hospital & Paracelsus Medical University; Institute for Pathology, Nuremberg General Hospital & Paracelsus Medical University; Department of Pediatrics, Nuremberg General Hospital & Paracelsus Medical University; Department of Gynecology and Obstetrics, Nuremberg General Hospital & Paracelsus Medical University; Institute for Clinical Chemistry, Laboratory Medicine and Transfusion Medicine, Nuremberg General Hospital & Paracelsus Medical University

**Keywords:** Cystathionine-γ-lyase, Cystathionine-β-synthase, oxidative stress, heme-oxygenase 1, homocysteine

## Abstract

Pre-eclampsia (PE) is a hypertensive pregnancy complication. Oxidative stress is hypothesized to contribute to the pathophysiology of PE. Both the hydrogen sulfide (H_2_S) and oxytocin (OT) systems might play a role in the pathophysiology of PE, liked to their anti-oxidant and hypotensive effects. Thus, the role of the interaction of the OT and H_2_S systems in the context of PE was further elucidated in the present clinical case control study “NU-HOPE” (**N**ürnberg-**U**lm: The role of **H**_2_S and **O**xytocin Receptor in **P**re-**E**clampsia; ethical approval by the Landesärztekammer Bayern, file number 19033, 29^th^ August 2019), comparing uncomplicated pregnancies, early-onset PE (ePE, onset <34 weeks gestational age) and late-onset PE (lPE, onset >34 weeks gestational age). Routine clinical data, serum H_2_S and homocysteine levels, tissue protein expression as well as nitrotyrosine formation were determined. The main findings were *(i)* unchanged plasma sulfide levels *(ii)* significantly elevated homocysteine levels in ePE, but not lPE, *(iii)* significantly elevated expression of H_2_S enzymes and OT receptor in the placenta in lPE, *(iv)* significantly elevated nitrotyrosine formation in the lPE myometrium. Taken together, these findings suggest a role for the interaction of the endogenous H_2_S- and OT/OTR systems in the pathophysiology of pre-eclampsia, possibly linked to impaired antioxidant protection.

## Introduction

Pre-eclampsia (PE) is one of the most common complications of pregnancy, the diagnostic criteria being: sudden onset of arterial hypertension during pregnancy (>20 weeks gestation) in combination with at least one other organ-specific complication, e.g., proteinuria and/or maternal liver dysfunction [1]. The pathophysiological mechanisms are not entirely clear, and the only causal therapy for PE currently available is the delivery of the baby [1]. Hypotheses on possible mechanisms for the development of PE comprise placental dysfunction or abnormal placentation [2], inflammatory processes [3], oxidative stress [3] and/or an imbalance in angiogenic factors [4].

Both the endogenously released gaseous signaling molecule hydrogen sulfide (H_2_S) as well as the neuropeptide oxytocin (OT, also known as the “cuddling hormone”) share anti-oxidant and anti-inflammatory effects [5]. H_2_S, a freely diffusible gas and thus acting without a receptor, is endogenously released by cystathionine-γ-lyase (CSE), cystathionine-β-synthase (CBS) and, to a lesser degree, 3-mercaptopyruvate-sulfurtransferase [6]. Oxytocin is mainly known for its role in parturition, social bonding and lactation, mostly acting through the activation of oxytocin receptor (OTR). Besides these roles, OT and OTR are also expressed in the cardiovascular system and implicated in the regulation of heart rate and blood pressure, with primarily hypotensive effects [7]. Similarly, genetic deletion of CSE, the main endogenous H_2_S-releasing enzyme in the cardiovascular system, is associated with a hypertensive phenotype in mice [8]. In turn, chronic hypertension as a pre-existing medical condition in the expecting mother is a significant risk factor for the development of PE [9]. In addition, the hemodynamic reaction of pre-eclamptic pregnant women has been reported to be less predictable and more heterogeneous compared to healthy pregnant women [10]. Thus, it is tempting to speculate, that the H_2_S- and OT-systems play a role in the underlying molecular mechanisms of PE. In myocardial samples from animal models of trauma, an interaction of both systems has been reported [11–14]. In fact, the interaction of H_2_S and OT/OTR has been proposed to be mediated by downstream signaling of the gaseous signaling molecule nitric oxide (NO) [15]. An interaction and reciprocal regulation of not only H_2_S and NO, but also of another gaseous mediator, carbon monoxide (CO), in context of the cardiovascular system and regulation of blood pressure is well established in the literature [16]. Moreover, a regulatory effect of CSE and CBS on OTR expression in the pregnant myometrium has been reported by You et al.: with the onset of labor, myometrial CSE and CBS are down-regulated, which is associated with an increase in contraction-associated proteins such as OTR [17]. However, their respective roles in the placenta might be more relevant for the pathophysiology of pre-eclampsia. For OT and OTR it has been reported that both their expressions increase with gestational age in the placenta [18]. OT mediates contraction of the myometrium, constriction of myometrial arteries and vaso-constriction in the chorionic plate, but not villous arteries [19]. A reduced OTR mRNA and protein expression has been reported in the placental vasculature of PE patients [20]. Surprisingly, however, very limited data on the role of placental OT/OTR in PE are available in the literature.

For the role of H_2_S and its endogenous enzymes in the placenta in PE, more literature reports are available. A reduction of CSE protein in the placenta of PE patients, along with a reduction of plasma sulfide levels, has been observed [21]. In contrast, Possomato-Vieira et al. report elevated plasma H_2_S in pre-eclamptic patients compared to healthy pregnant women [22]. Another study reported unchanged CBS and CSE protein in the placenta of PE patients [23]. Cindrova-Davies et al. observed no differences between control and PE patients in placental CBS protein. CSE protein in PE placentas with abnormal blood flow through the umbilical artery was reduced and CSE protein in PE placentas with normal umbilical blood flow was elevated compared to control tissue [24]. Hence, in summary, literature reports are contradictory, and there is no consistent pattern for the regulation of H_2_S enzymes during pre-eclampsia.

Given the abovementioned possible OT/H_2_S interaction and the scarce data available on OT/OTR in PE, the aim of this study was to further elucidate the role of OT/OTR and the interaction with the endogenous H_2_S system in the context of pre-eclampsia.

## Material and Methods

This case control study „NU-HOPE” (**N**ürnberg-**U**lm: The role of **H**_2_S and **O**xytocin Receptor in **P**re-**E**clampsia) was a collaborative effort of Ulm University Medical Center (Institute for Anesthesiological Pathophysiology and Process Engineering) and Nuremberg Hospital (Clinic for Psychosomatic Medicine and Psychotherapy; Clinic for Gynecology and Obstetrics; Institute for Clinical Chemistry, Laboratory Medicine and Transfusion Medicine). Ethical approval with adherence to the Declaration of Helsinki was granted by the Landesärztekammer Bayern (file number 19033, 29^th^ August 2019). Patient recruitment was started in October 2019 and concluded in March 2020 at the Clinic for Gynecology and Obstetrics at Nuremberg South Hospital. The processing and analysis of the collected blood samples and data were performed at the Clinic for Psychosomatic Medicine and Psychotherapy and at the Institute for Clinical Chemistry, Laboratory Medicine and Transfusion Medicine in Nuremberg. Samples for sulfide detection, isolated PBMCs and tissue samples were transferred to the Institute for Anesthesiological Pathophysiology and Process Engineering at Ulm University Medical Center.

After written informed consent had been obtained, pregnant women scheduled for elective cesarean section for a variety of reasons (breach presentation, maternal risk factors) were recruited for the control group (N=54). For the case group, pregnant women with diagnosis of pre-eclampsia were recruited (initially N=49). According to current clinical guidelines for pre-eclampsia, inclusion criteria for the case group were hypertension (systolic/diastolic blood pressure ≥ 140/90 mmHg) and newly emerging organ-specific manifestation (i.e., kidney: proteinuria in 24h-urine ≥ 300 mg *AND/OR* liver: ≥ two-fold elevation of alanine-aminotransferase (ALT) and aspartate-aminotransferase (AST) *AND/OR* hematologic system: thrombocytopenia < 100 Gpt/*AND/OR* central nervous system: presentation with tonic-clonic seizure *OR* placenta intrauterine growth restriction (IUGR), small for gestational age (SGA)). Exclusion criteria were lacking skills of German, twin pregnancies, Type I or II diabetes mellitus, chronic-inflammatory diseases. One patient from the control group developed pre-eclampsia at a later time-point and, thus, was re-assigned to the case group. In three patients from the case group, the pre-eclampsia diagnosis was not confirmed during a later medical examination, so that they were dropped from the study. Patients who failed to show up for follow-up appointments after recruiting were equally dropped from the study. The final N per group was N=51 in the control group and N=44 in the case group. The case group was further divided into patients with early (<34 weeks pregnant; N=8) and late (≥34 weeks pregnant; N=36) onset pre-eclampsia [1]. The trial structure is visualized in Figure 1A.

**Figure 1:**
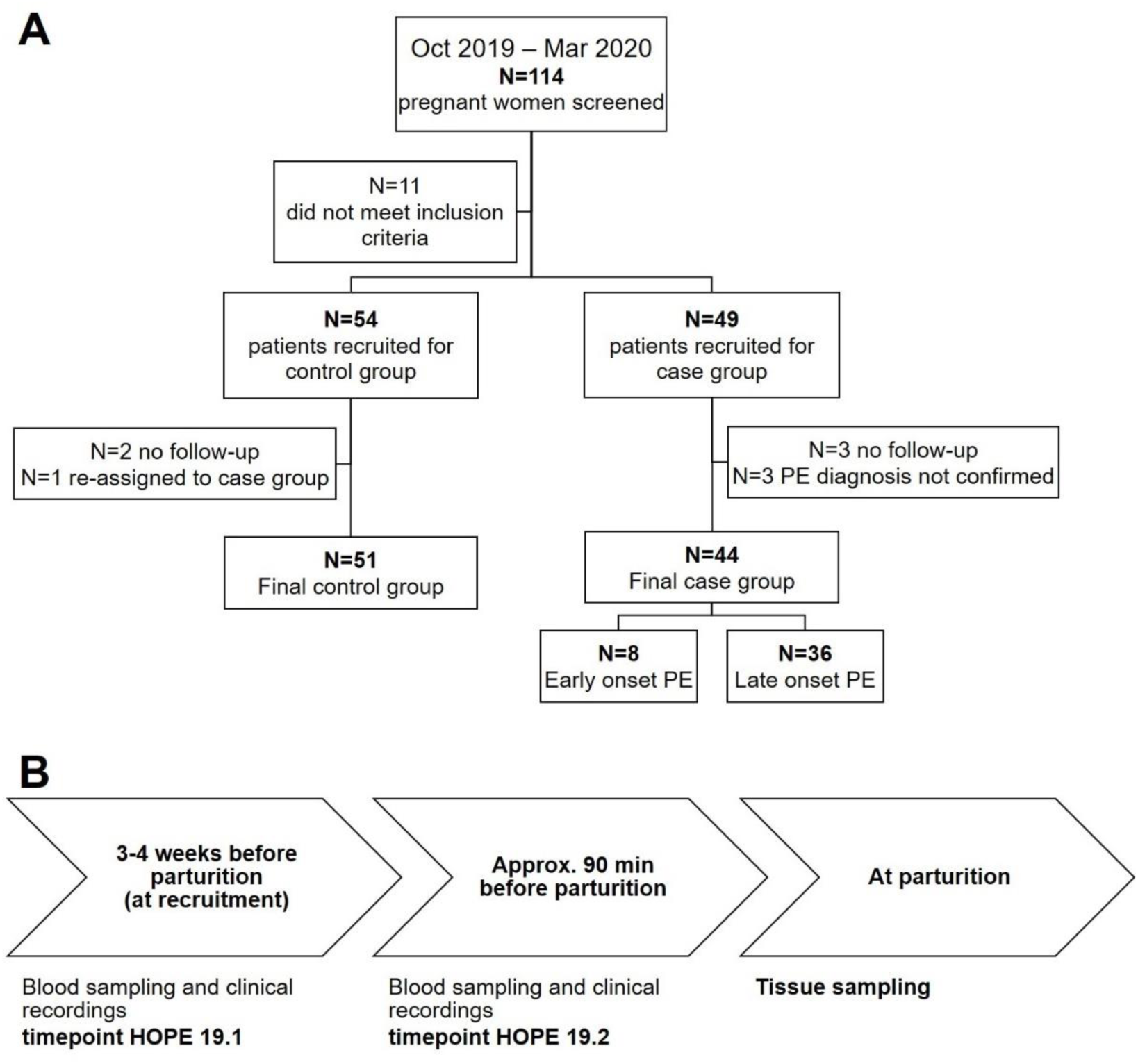
Recruitment and sampling procedures. **A** Trial profile. Over a period of 6 months, all pregnant women, which had an appointment for a scheduled cesarean section and all pregnant women with suspected pre-eclampsia were approached and all patients fulfilling the inclusion critera were recruited for the study after written informed consent. Some patients dropped out of the study, because they did not show up for follow-up appointments, one control patient had to be reassigned to the case group because of developing pre-eclampsia. Three patients of the case group were dropped from the study, because the initial diagnosis of pre-eclampsia was not confirmed at a later timepoint. The case group was further divided into early- and late-onset pre-eclampsia. **B** Time course of the study.

### Sampling procedures

A timeline for the sampling procedures is given in Figure 1B. Three to four weeks (timepoint HOPE 19.1) before the calculated date of parturition, blood was sampled from control patients for PBMC isolation, measurement of sulfide levels and analysis of various clinical parameters (AST, ALT, protein, albumin, urea, creatinine, bilirubin, CRP, estimated glomerular filtration rate (eGFR), calcium, potassium, sodium, parameters of coagulation, cell counts). This timepoint varied in the case groups depending on urgency: blood sampling was performed as early as three to four weeks before, up until the date of parturition, when parturition was imminent at the date of recruitment. Patients of the pre-eclampsia group either had spontaneous births, or babies were delivered by cesarean section, depending on the severity of pre-eclampsia, the decision being made by the attending obstetrician. Patients of the control group were prepared in the delivery room approx. 90 min before the scheduled cesarean section. Shortly before parturition, another blood sampling in the fasted patients (timepoint HOPE 19.2) was performed to determine homocysteine, folic acid, vitamin B12, vitamin B6 levels.

Myometrial (only cesarean sections) and placental biopsies (all patients) were sampled shortly after parturition.

### Clinical Parameters

All routine biological parameters (AST, ALT, protein, folic acid, Vit B6 and Vit B12, creatinine, thrombocyte counts) were determined by the Institute for Clinical Chemistry, Laboratory Medicine and Transfusion Medicine of Nuremberg South Hospital. For the determination of homocysteine levels, blood was sampled in the specified monovette “Homocystein Z-Gel” after 12h of fasting. The sample was centrifuged, and the supernatant was measured *via* a chemiluminescent microparticle assay with the Architect 1000 I SR analyzer (Abbott, Wiesbaden, Germany).

### Sulfide Measurements

Sulfide measurements were performed with a well-established technique developed in house [25]. Blood was sampled into a 2.7 mL Li-Heparin Monovette. 3 times 100 µL (to be able to measure all samples in triplicates) were centrifuged for 3 min at room temperature to gain a plasma sample. Sulfide contained in the blood plasma was immediately derivatized into the Di-(pentafluorbenzyle-)-derivate. 100 µL of sample (blood, plasma or calibration solution) were added to the derivatization mixture (400 µL internal standard (2 µg/mL 1,3,5-tribromebenzol in isooctane) + 200 µL alkylation reagent (10 µL/ml pentafluorobenzylbromide in isooctane) + 400 µL phase transfer catalyzer (2 mg/mL benzyldimethyltetradecylammoniumchlorise in *aqua bidest* saturated with sodium tetraborate). The mixture was vigorously vortexed for 1 min, 400 µL of water saturated with potassium dihydrogenphosphate were added, vortexed again for 10 sec and centrifuged for 2 min at 13000 rpm. The lower aqueos phase was frozen out at -80 °C, and the upper organic phase was transferred to a fresh vial. The derivatized samples were stored at -80 °C and transferred to the Institute for Anesthesiological Pathophysiology and Process Engineering at Ulm University Medical Center to determine sulfide levels *via* gas chromatography and mass spectrometry (GC/MS, Agilent 6890/5973 GC/MS system with capillary column MN Optima 5-MS (12m x 0.2 mm; 0.35 µm film thickness), 5 psi, and helium as a carrier gas). Calibration samples (0.1 µM – 4 µM) were generated by dissolving sodium sulfide in 0.01 M sodium tetraborate. Signal ratios (sulfide/internal standard) were correlated with concentration ratios (sulfide/internal standard) to obtain calibration curves. Injector temperature was 250 °C and 2 µL of samples were injected at an oven temperature of 80 °C. After 1 min, heating started with 25°/min up to 200 °C and 50°/min to 300 °C. The m/z values of 313.7 (internal standard, retention time 4.6 min) and 394.0 (sulfide derivate, retention time 5.7 min) were recorded after electron impact ionization at 70 eV.

### Isolation of PBMC

27 mL blood were sampled into 3×9 ml EDTA monovettes and, after 1:1 dilution with Dulbecco’s Phosphate Buffered Saline, layered on top of 1,119 g/mL and 1.077 g/mL Ficoll gradient. The gradients were centrifuged for 20 min at room temperature (2173 rpm). The resulting Buffy-ring of PBMC was transferred to a fresh tube, filled up with DPBS and centrifuged again at 4 °C for 10 min (1797 rpm). Erythrocytes were lysed by adding aqua bidest., subsequent neutralization with 10X DPBS (10:1), filling up with 1X DPBS and centrifuged for 7 min at 4 °C (1797 rpm). The supernatant was discarded and the cell pellet was washed with DPBS and centrifuged again at 4 °C for 7 min (1797 rpm). Cells were counted with the help of a Neubauer chamber, re-pelleted (7 min centrifugation at 1797 rpm), resuspended in DPBS and aliquoted (10 Mio cells). Cells were centrifuged again for 3 times 10 sec at 13000 rpm to remove the DPBS and dry the pellet. Cell pellets were stored at -80 °C.

### Tissue processing

Uterine biopsies were taken from the upper border of the cesarean section related incision (0.5-1 cm^3^) and contained endometrium, myometrium and serosa. One part of the uterine biopsy as well as a placenta sample containing fetal and maternal parts were snap frozen in liquid nitrogen and stored at -80 °C. The other part of the uterine biopsy as well as a 2 cm^3^ piece of placenta with maternal and fetal parts were immediately fixed in formalin. Two hours later, these samples were cut into 4 mm sections, transferred to embedding cassettes and fixed in formalin for another 48h. Dehydration and embedding in paraffin was performed at the Institute for Pathology at Nuremberg North Hospital.

### Immunohistochemistry

Immunohistochemical analysis was used to assess levels of OTR, the H_2_S-producing enzymes CSE and CBS, and nitrotyrosine formation (a marker of oxidative and nitrosative stress [26]) in the myometrium and intervillous space of the placenta. Immunohistochemistry was chosen for several reasons: (i) it is widely acknowledged in the literature that densitometric analysis of colorimetric immunohistochemical staining provides comparable reliability to Western blotting for quantifying protein levels [27]; (ii) significant correlations have been observed between densitometric values and those obtained from Western blotting analysis [28]; and (iii) in contrast to Western blotting, immunohistochemical analysis of tissue enables the identification of spatial distribution and protein expression in specific cell types within the tissue sample. Immunohistochemistry was performed following the methodology outlined previously [29]. Formalin-fixed and paraffin embedded tissue sections of 3–5 μm thickness underwent deparaffinization using xylene, followed by rehydration through a sequential series of ethanol and deionized water. Heat-induced antigen retrieval was conducted in citrate solution (pH 6). Subsequently, slides were blocked with 10% goat serum (Jackson ImmunoResearch Laboratories, Inc., West Grove, PA, UK) prior to incubation with the following primary antibodies: H_2_S-producing enzymes anti-CBS (Protein Tech, Manchester, UK, 14787-1-AP, RRID: AB_2070970), anti-CTH (Abnova, H00001491-M03, RRID:AB_489881), anti-OT-R (Protein Tech, 2304523045-1-AP, RRID: AB_2827435), and anti-nitrotyrosine (Merck SA, an affiliate of Merck KGaA, Darmstadt, Germany, ab5411, RRID:AB_177459). The optimal dilution for all primary antibodies was titrated to optimal concentrations guided by the manufacturer’s recommendations. Primary antibodies were detected using the Dako REAL detection system (anti-mouse, anti-rabbit, alkaline phosphatase-conjugated), which uses a red chromogen as substrate. Slides were counterstained with hematoxylin (Sigma, St. Louis, MO, USA). The slides were examined using a Zeiss Axio Imager A1 microscope and quantitative analysis was conducted on 800,000 μm^2^ sections utilizing the Zen Image Analysis Software (Zeiss, Oberkochen, Germany). The findings are presented as the percentage of positively stained area relative to the total area [30].

### Western blotting

Immunoblotting for CSE, OTR, and heme oxygenase-1 (as a marker of the antioxidant response) was performed as described previously [31,32]. Tissue samples were homogenized and lysed in lysis buffer. Cell pellets were re-suspended, lysed on ice and centrifuged. The supernatant (protein extract) was stored at -80°C. protein concentrations were determined, and equal total protein aliquots (20–60 μg) were separated by SDS-PAGE and transferred by Western blotting. After blocking, the membranes were incubated with commercially available primary antibodies (anti-CSE Protein Tech, 12217-1-AP, RRID: AB_2087497, anti-OTR Protein Tech, 2304523045-1-AP, RRID: AB_2827435, anti-HO-1; Abcam, ab52947,

RRID:AB_880536). Primary antibodies were detected by using horseradish peroxidase-conjugated secondary anti-rabbit IgG antibody (Cell Signaling #7074, RRID: AB_2099233). b-actin (Santa Cruz Biotechnology sc-1615 RRID: AB_880536) served as a loading control. The membranes were subjected to chemo-luminescence using SuperSignalWest Femto Maximum Sensitivity Substrate (Thermo Fisher Scientific). Exposed films were scanned, and intensity of immune-reactivity was densitometrically measured using NIH Image J software (http://rsb.info.nih.gov/nih-image). Bands specific for HO-1 (37 kD, according to antibody manufacturer), CSE (44 kD, according to antibody manufacturer), OTR (two bands, 46 kD and 67 kD, according to antibody manufacturer and [33]) and Actin (45 kD, according to antibody manufacturer) were analyzed. All immuno-blots were repeated twice. For normalization to protein loading, the band intensities for the proteins of interest were related to actin as loading control. For comparison between individual gels, each sample was related to HeLa cell extracts (tissue samples) or PBMC of a healthy volunteer (PBMC samples). Results are presented as densitometric sum.

### Statistics

Statistical analysis was performed with GraphPad Prism 8. All data are presented as median [lower quartile; upper quartile], unless otherwise stated. If testing for normal distribution was passed, intergroup differences were assessed via one-way ANOVA with a post hoc Tukey test for multiple comparisons. Data with no normal distribution were further analyzed with Kruskal-Wallis ANOVA and a post hoc Dunn’s test for multiple comparisons. A Mann-Whitney-Test was used when the data were only available for two groups. Data sets with matched values (e.g., blood pressure, Apgar) were analyzed with a two-way ANOVA and post hoc Sidak’s test for multiple comparisons. Linear correlations were tested by calculating the correlation coefficient according to Pearson. Linear modelling was performed with the following equation: y=ax+b.

## Results

### Patient demographics and clinical parameters

Demographic and clinical patient characteristics are represented in Table 1. There were no significant differences between the study groups in the demographic parameters such as age, height and weight. 39% of mothers in the late-onset pre-eclampsia (lPE) group had a vaginal delivery, while there were none in the early-onset pre-eclampsia (ePE). In accordance with the diagnostic criteria for pre-eclampsia (PE), both case groups had a significantly elevated systolic and diastolic blood pressure at hospital admission; in addition, the systolic pressure was significantly higher in the ePE group compared to lPE. Furthermore, PE groups had higher total plasma protein compared to controls and 7 out of 8 and 24 out of 29 patients in the ePE and lPE met the threshold of > 300 mg/24h in their urine, i.e., fulfilled the diagnostic criterium for proteinuria. Proteinuria was significantly more severe in ePE than lPE. Furthermore, creatinine levels and liver enzymes (AST, ALT) were elevated in the PE groups compared to controls. There were no cases of thrombocytopenia and no intergroup differences hemoglobin, folic acid, vitamin B6 and vitamin B12.

**Table 1:**
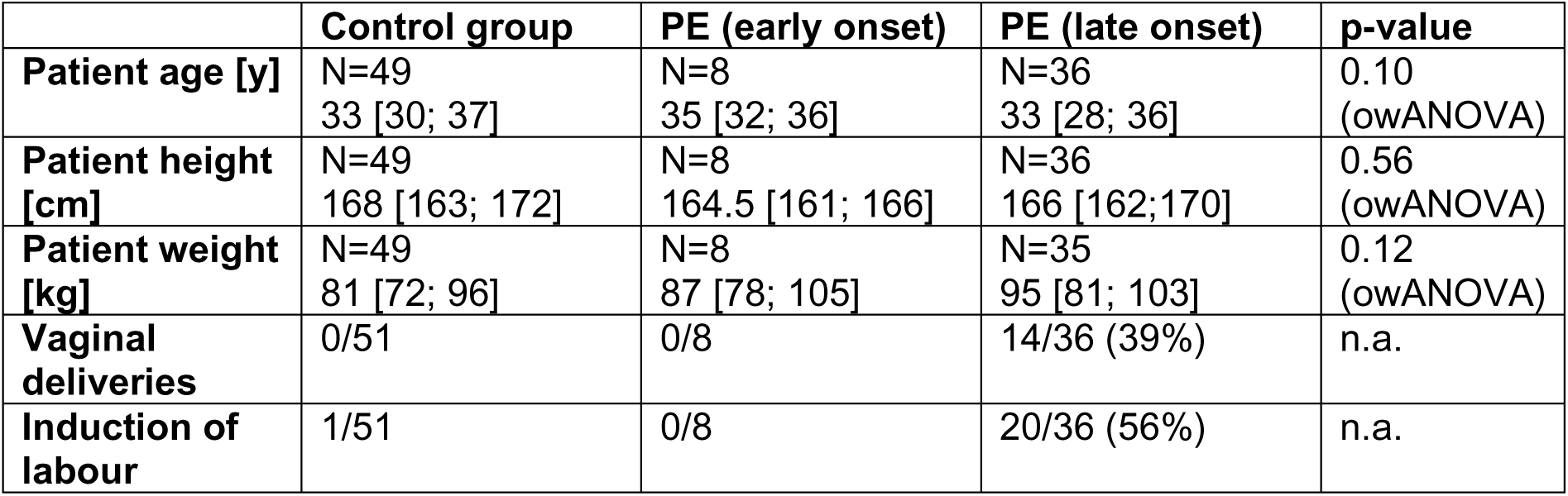
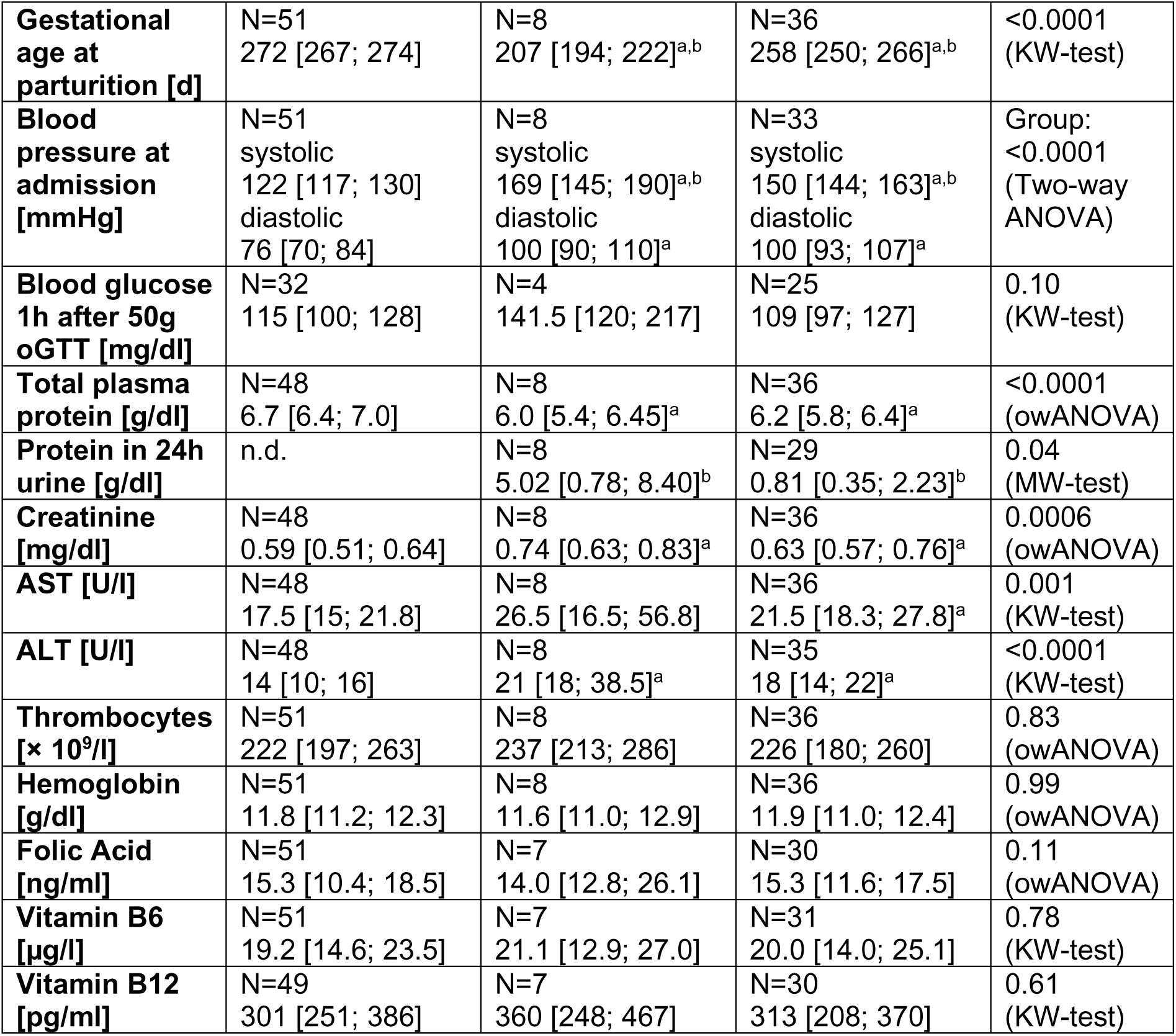
Demographic and clinical parameters of the study groups. Data are median [25% quartile; 75% quartile]. a significant vs. Control, b significant vs. other pre-eclampsia group. Pre-eclampsia (PE), one-way analysis of variance (owANOVA), Kruskal-Wallis-test (KW-test), Mann-Whitney-test (MW-test), Oral Glucose Tolerance Test (oGTT)

|  | Control group | PE (early onset) | PE (late onset) | p-value |
| --- | --- | --- | --- | --- |
| **Patient age [y]** | N=49<br>33 [30; 37] | N=8<br>35 [32; 36] | N=36<br>33 [28; 36] | 0.10<br>(owANOVA) |
| **Patient height [cm]** | N=49<br>168 [163; 172] | N=8<br>164.5 [161; 166] | N=36<br>166 [162; 170] | 0.56<br>(owANOVA) |
| **Patient weight [kg]** | N=49<br>81 [72; 96] | N=8<br>87 [78; 105] | N=35<br>95 [81; 103] | 0.12<br>(owANOVA) |
| **Vaginal deliveries** | 0/51 | 0/8 | 14/36 (39%) | n.a. |
| **Induction of labour** | 1/51 | 0/8 | 20/36 (56%) | n.a. |
| **Gestational age at parturition [d]** | N=51<br>272 [267; 274] | N=8<br>207 [194; 222]<sup>a,b</sup> | N=36<br>258 [250; 266]<sup>a,b</sup> | <0.0001<br>(KW-test) |
| **Blood pressure at admission [mmHg]** | N=51<br>systolic<br>122 [117; 130]<br>diastolic<br>76 [70; 84] | N=8<br>systolic<br>169 [145; 190]<sup>a,b</sup><br>diastolic<br>100 [90; 110]<sup>a</sup> | N=33<br>systolic<br>150 [144; 163]<sup>a,b</sup><br>diastolic<br>100 [93; 107]<sup>a</sup> | Group:<br><0.0001<br>(Two-way ANOVA) |
| **Blood glucose 1h after 50g oGTT [mg/dl]** | N=32<br>115 [100; 128] | N=4<br>141.5 [120; 217] | N=25<br>109 [97; 127] | 0.10<br>(KW-test) |
| **Total plasma protein [g/dl]** | N=48<br>6.7 [6.4; 7.0] | N=8<br>6.0 [5.4; 6.45]<sup>a</sup> | N=36<br>6.2 [5.8; 6.4]<sup>a</sup> | <0.0001<br>(owANOVA) |
| **Protein in 24h urine [g/dl]** | n.d. | N=8<br>5.02 [0.78; 8.40]<sup>b</sup> | N=29<br>0.81 [0.35; 2.23]<sup>b</sup> | 0.04<br>(MW-test) |
| **Creatinine [mg/dl]** | N=48<br>0.59 [0.51; 0.64] | N=8<br>0.74 [0.63; 0.83]<sup>a</sup> | N=36<br>0.63 [0.57; 0.76]<sup>a</sup> | 0.0006<br>(owANOVA) |
| **AST [U/l]** | N=48<br>17.5 [15; 21.8] | N=8<br>26.5 [16.5; 56.8] | N=36<br>21.5 [18.3; 27.8]<sup>a</sup> | 0.001<br>(KW-test) |
| **ALT [U/l]** | N=48<br>14 [10; 16] | N=8<br>21 [18; 38.5]<sup>a</sup> | N=35<br>18 [14; 22]<sup>a</sup> | <0.0001<br>(KW-test) |
| **Thrombocytes [$\times 10^9/l$]** | N=51<br>222 [197; 263] | N=8<br>237 [213; 286] | N=36<br>226 [180; 260] | 0.83<br>(owANOVA) |
| **Hemoglobin [g/dl]** | N=51<br>11.8 [11.2; 12.3] | N=8<br>11.6 [11.0; 12.9] | N=36<br>11.9 [11.0; 12.4] | 0.99<br>(owANOVA) |
| **Folic Acid [ng/ml]** | N=51<br>15.3 [10.4; 18.5] | N=7<br>14.0 [12.8; 26.1] | N=30<br>15.3 [11.6; 17.5] | 0.11<br>(owANOVA) |
| **Vitamin B6 [μg/l]** | N=51<br>19.2 [14.6; 23.5] | N=7<br>21.1 [12.9; 27.0] | N=31<br>20.0 [14.0; 25.1] | 0.78<br>(KW-test) |
| **Vitamin B12 [pg/ml]** | N=49<br>301 [251; 386] | N=7<br>360 [248; 467] | N=30<br>313 [208; 370] | 0.61<br>(KW-test) |

The babies of both pre-eclampsia groups were born at a significantly earlier gestational age (see Table 1), and the newborns had significantly lower birth weights when compared to the control group (see Table 2). The control group had a slightly higher fraction of female newborns, whereas the newborn sex distribution was more equal in the pre-eclampsia groups. Apgar scores were rising in all groups over time but the significantly lowest values occured in ePE newborns compared to both other groups (Two-way ANOVA p<0.0001 for factor time and p<0.001 for factor group).

**Table 2:**
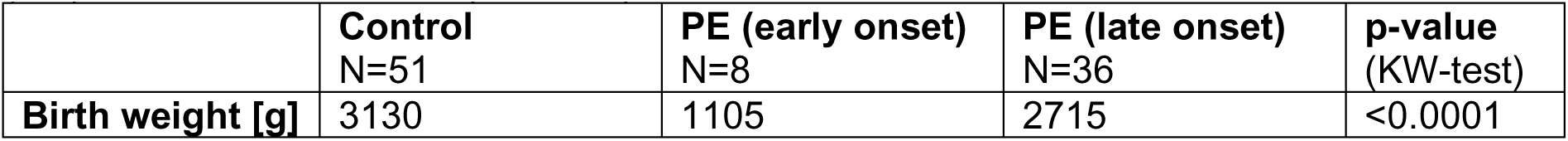
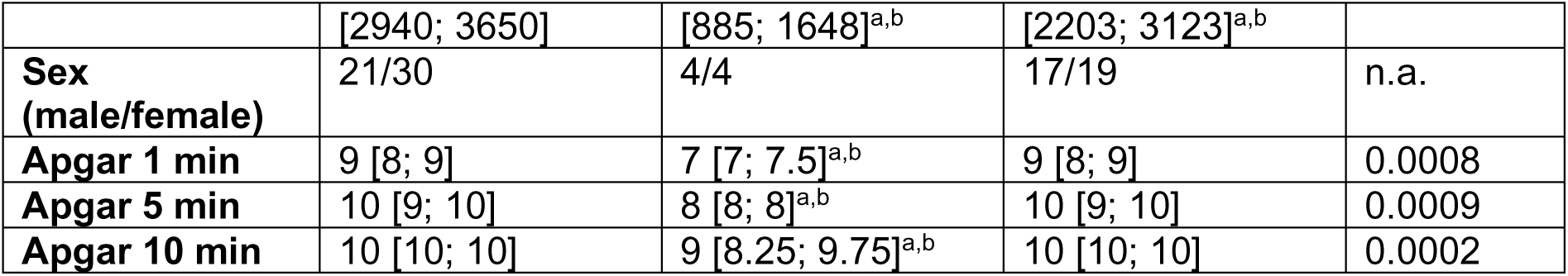
Outcome of the newborns. Data are median [25% quartile; 75% quartile]. a significant vs. Control, b significant vs. other pre-eclampsia group. Pre-eclampsia (PE), Kruskal-Wallis-test (KW-test)

|  | **Control**<br>N=51 | **PE (early onset)**<br>N=8 | **PE (late onset)**<br>N=36 | **p-value**<br>(KW-test) |
| --- | --- | --- | --- | --- |
| **Birth weight [g]** | 3130 | 1105 | 2715 | <0.0001 |
|  | [2940; 3650] | [885; 1648]<sup>a,b</sup> | [2203; 3123]<sup>a,b</sup> |  |
| **Sex (male/female)** | 21/30 | 4/4 | 17/19 | n.a. |
| **Apgar 1 min** | 9 [8; 9] | 7 [7; 7.5]<sup>a,b</sup> | 9 [8; 9] | 0.0008 |
| **Apgar 5 min** | 10 [9; 10] | 8 [8; 8]<sup>a,b</sup> | 10 [9; 10] | 0.0009 |
| **Apgar 10 min** | 10 [10; 10] | 9 [8.25; 9.75]<sup>a,b</sup> | 10 [10; 10] | 0.0002 |

*Biological correlates of the Hydrogen sulfide (H_2_S)- and Oxytocin (OT)-systems* Blood levels of homocysteine, a substrate for the H_2_S-enzyme cystathionine-β-synthase, were significantly elevated in ePE compared to control (see Figure 2). There was no statistically significant difference between lPE and ePE, suggesting that homocysteine levels in this group were slightly elevated as well, though not significantly different from control (see Figure 2). Plasma sulfide levels did not differ between the groups (see Figure 2). The N in the PE-groups was rather low for this analysis (see Figure 2). Due to the high workload for this analysis, the required sample workup was stopped pre-maturely after it became clear that it was unlikely that there would be a significant intergroup difference. Hence, the N for the PE groups (see Figure 2) in this analysis is lower than the total number of recruited patients.

**Figure 2:**
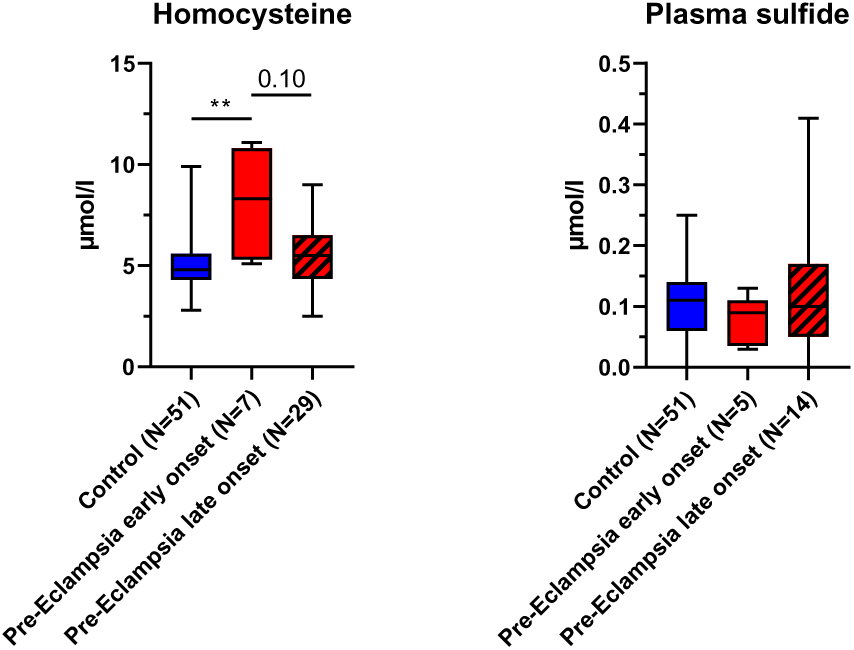
Homocysteine and sulfide levels. Homocysteine p=0.008 (Kruskal-Wallis-test); ** p<0.01. Plasma sulfide p=0.59 (One-way ANOVA).

In the placental intervillous layer, CSE expression was mostly present in the endothelium of fetal capillaries and villous tissue (see exemplary pictures, Figure 3 top row). In control tissues, CBS expression was limited to the villous trophoblast layer, whereas in PE placentas, CBS was additionally expressed in the maternal and fetal capillary blood (see exemplary pictures, Figure 3, second row). In lPE, CBS was also present in the villous tissue (see exemplary pictures, Figure 3, second row). OTR was always present in the fetal capillariesand villous tissue. In lPE, the villous trophoblast layer was additionally positive for OTR (see exemplary pictures, Figure 3, third row). Nitrotyrosine formation, if present, was found in the villous tissue and fetal capillary blood and endothelium (exemplary pictures, Figure 3, bottom row). CBS and oxytocin receptor (OTR) expression were significantly elevated in placentas from lPE patients in comparison to the control group (see Figure 3, right column). The expression of cystathionine-γ-lyase was also elevated in lPE patients compared to controls, though just missing statistical significance (p=0.06) (see Figure 3, right column). There was a trend towards higher placental nitrotyrosine formation in these patients as well (see Figure 3, right column). There were no differences in placental proteins between controls and ePE patients (see Figure 3, right column).

**Figure 3:**
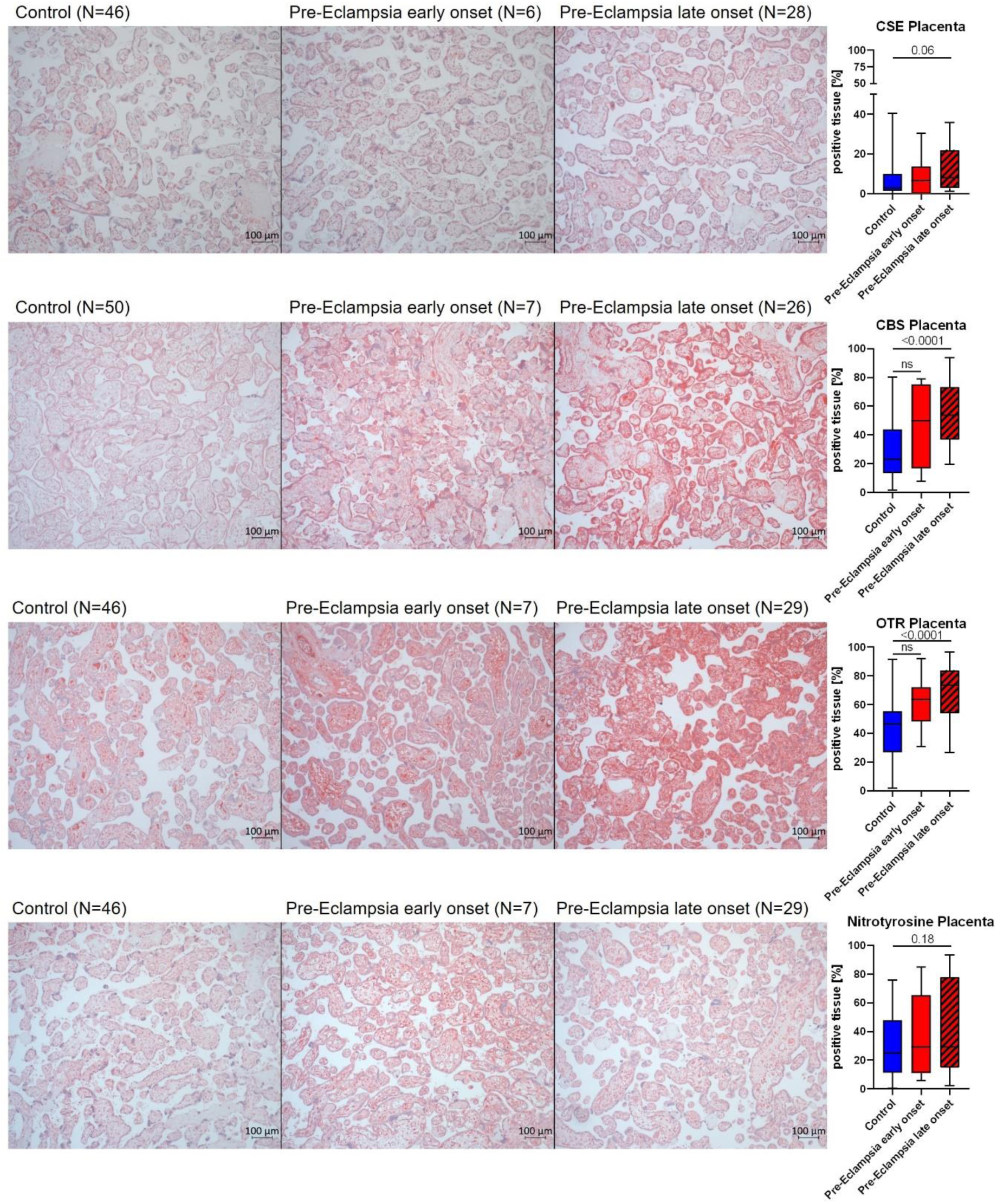
Representative pictures with the IHC target stained in red (left) and quantification (% positive tissue, right) of protein expression in the placenta. CSE, CBS and Nitrotyrosine were statistically analyzed with Kruskal-Wallis-test. OTR was statistically analyzed with one-way ANOVA. The N per group is subject to variation, since some samples could not be used for the final analysis due to technical reasons (i.e., compromised tissue, compromised IHC signal). This was not consistent throughout the groups and/or for specific target protein, which is why the N for the individual analyses is given within the figure.

Muscle cells of the uterine myometrium and capillary smooth muscle cells were highly positive for CSE, whereas layers of connective tissue in between the muscle fibers were negative (exemplary pictures, Figure 4 top row). CBS expression was very limited and localized to small blood vessels and myometrial muscle cells (exemplary pictures, Figure 4 second row). OTR was expressed in the uterine myometrium, as well as the endothelium and lumen of small blood vessels (exemplary pictures, Figure 4 third row). Nitrotyrosine formation, if present, was localized to muscle cells and blood (exemplary pictures, Figure 4 bottom row). Nitrotyrosine formation in the uterus of lPE patients was significantly more pronounced than in ePE and controls (see Figure 4). There were no intergroup differences for the expression of CSE, CBS and OTR in the uterus, except for a trend towards higher CBS expression in lPE just missing statistical significance (p=0.06) (see Figure 4). Neither the mode of delivery (vaginal vs. c-section) nor the fact if labour was induced in the patients or not was a relevant confounder for placental or uterine protein expression. This has been systematically analyzed for the lPE group, where there was a relevant N for both vaginal vs. c-section deliveries as well as induced births (data not shown).

**Figure 4:**
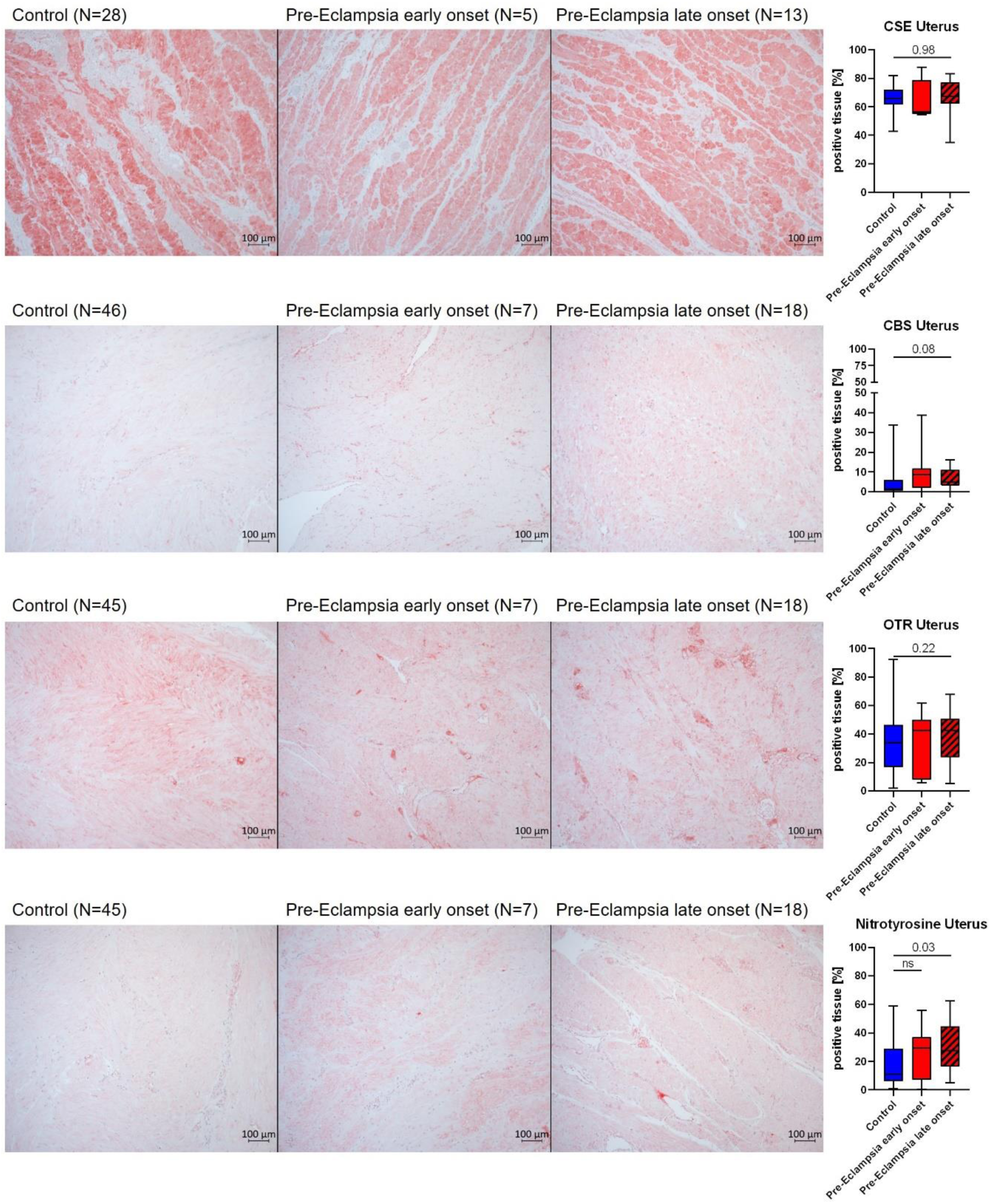
Representative pictures with the IHC target stained in red (left) and quantification (% positive tissue, right) of protein expression in the myometrium of uterine samples. CSE and OTR were statistically analyzed with one-way ANOVA. CBS and Nitrotyrosine were statistically analyzed with Kruskal-Wallis-test. The N per group is subject to variation, since some samples could not be used for the final analysis due to technical reasons (i.e., compromised tissue, compromised IHC signal). This was not consistent throughout the groups and/or for specific target protein, which is why the N for the individual analyses is given within the figure.

PBMC expression of CSE, OTR and heme oxygenase 1 (HO-1, as an endogenous producer of the gaseous mediator carbon monoxide and enzyme of the antioxidant defense) were analyzed *via* Western Blot. HO-1 expression was higher in both PE groups compared to control, however only statistically significant in lPE. The increase seemed even more pronounced in ePE patients, however most likely the low N in this group was the reason for the lack of statistical significance (see Figure 5). PBMC expression of CSE and OTR did not differ between groups. We had to use a different antibody for CSE for Western Blot than for IHC due to the fact that we were not able to establish a working Western Blot protocol with the antibody used for IHC. Thus, we decided to also analyze placental CSE expression *via* Western Blot confirming the IHC findings of a trend towards higher CSE expression in lPE patients compared to controls (see Figure 5). Furthermore, we also used Western Blot to determine placental HO-1 expression, which did not differ between groups.

**Figure 5:**
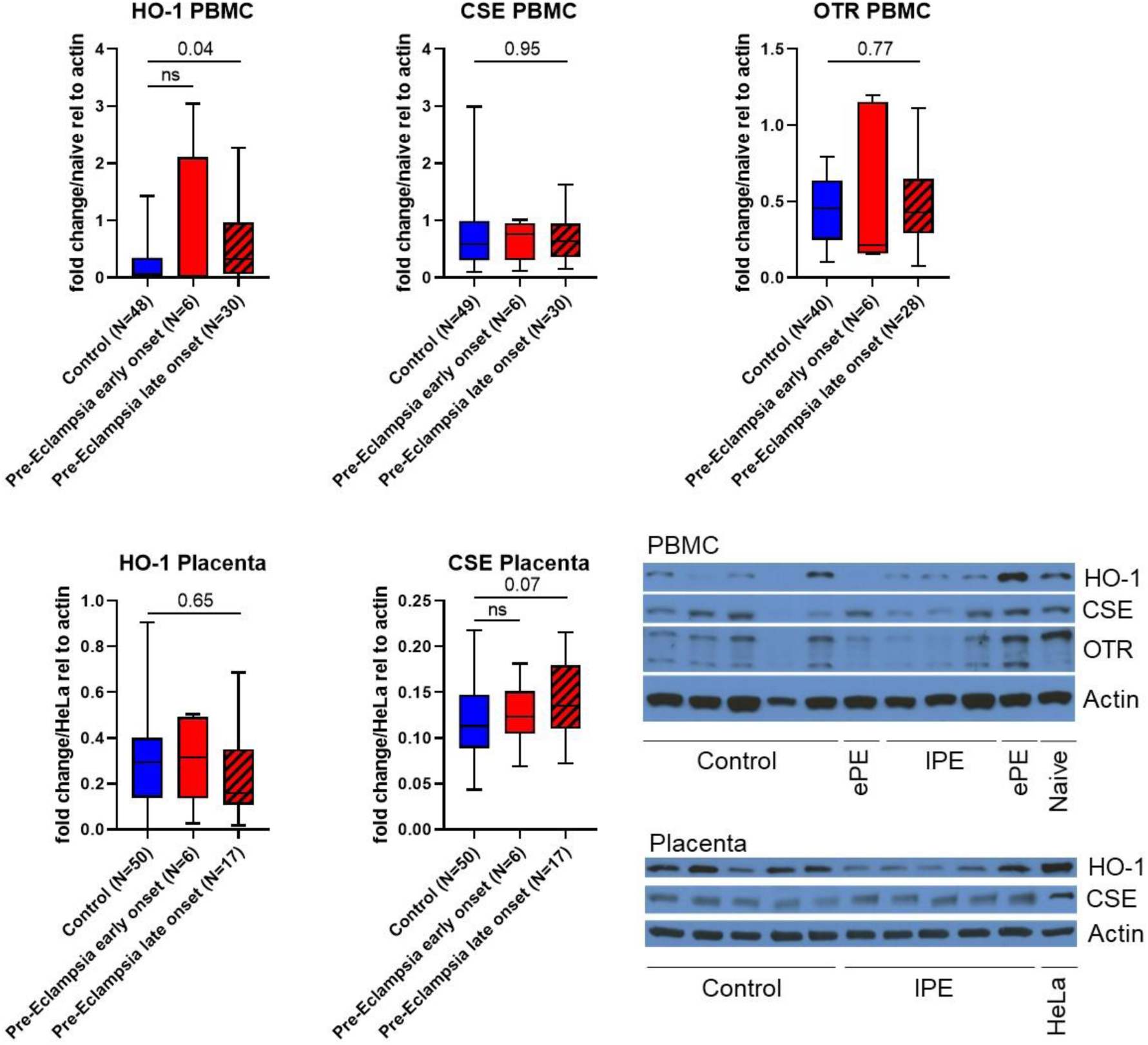
Quantification of protein expression via Western Blots. Top: HO-1, CSE and OTR expression in PBMC. PBMC from a healthy volunteer were used as naïve control. Bottom: HO-1 and CSE expression in the placenta. Protein extracts from HeLa cells were used as control. Bottom right: exemplary Western Blots. HO-1, CSE and OTR in PBMC were statistically analyzed with the Kruskal-Wallis-test. CSE and HO-1 in the placenta were statistically analyzed with One-way ANOVA. ePE = early onset pre-eclampsia, lPE = late onset pre-eclampsia.

Given the parallel effects of OT/OTR and H_2_S/CSE/CBS we also decided to analyze whether the expressions of these proteins were related with each other in the placenta. We were able to confirm a direct linear relationship between OTR and CBS as well as CSE, however the correlation with CBS was more robust than the one with CSE (see Figure 6 top row). In the uterus, there was a significant direct linear relationship between CBS and OTR expression and no significant correlation of CSE and OTR (see Figure 6 bottom row). Furthermore, we were interested if PBMC CSE or OTR expression might be a correlate for the protein expression in other, less accessible tissues (i.e., placenta and uterus). There were no significant relations between PBMC and placental/uterine CSE expression (p=0.35 and p=0.45 respectively, graphs not shown). There was no significant relation either between PBMC and placental OTR expression (see Figure 6. Bottom row), but there was a significant direct linear relationship between PBMC and uterine OTR expression (see Figure 6, bottom row).

**Figure 6:**
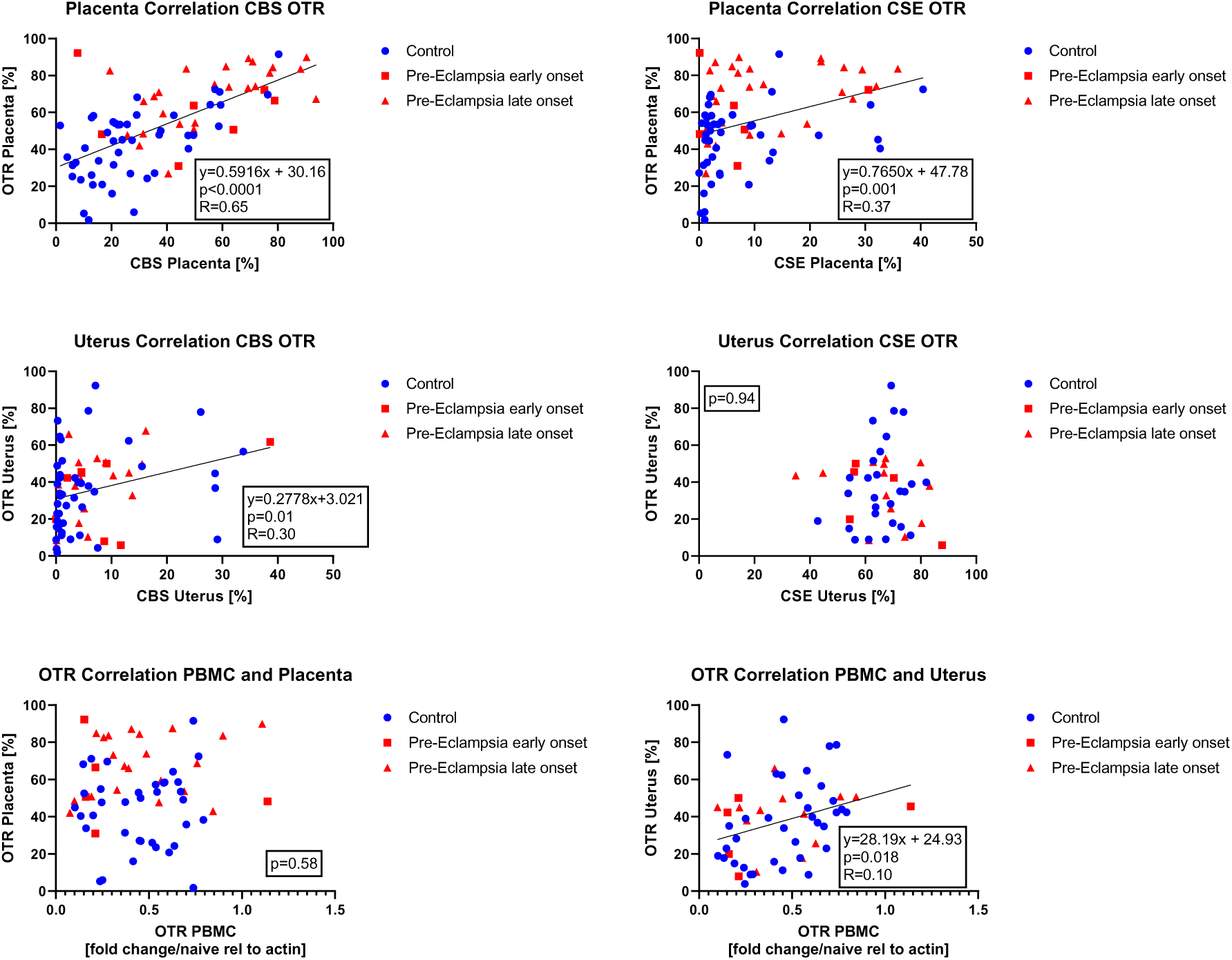
Correlations analysis (Pearson) of placental CBS and OTR expression with linear regression (top left), placental CSE and OTR expression with linear regression (top right), uterine CBS and OTR expression with linear regression (middle left), uterine CSE and OTR expression (middle right, no correlation), PBMC and placental OTR expression (bottom left, no correlation) and PBMC and uterine OTR expression with linear regression (bottom right).

## Discussion

The aim of this study was to further elucidate the role of the H_2_S- and OT-systems in pre-eclampsia (PE). The main findings were *(i)* unchanged plasma levels for sulfide between controls, early and late onset PE, *(ii)* significantly elevated homocysteine levels in early, but not late onset PE, *(iii)* significantly elevated CBS and OTR expression and slightly elevated CSE expression in the placenta in late-onset PE, *(iv)* significantly elevated nitrotyrosine formation in the uterine myometrium of late-onset PE patients, *(v)* elevated PBMC HO-1 expression in lPE with unchanged placental HO-1 expression, and *(vi)* significant direct relationships between placental OTR and CBS/CSE expression. Taken together, these findings suggest an interaction of the OTR and CSE/CBS in the placenta, associated with an elevation of these proteins during PE. In addition, PBMC protein expression is a very limited surrogate for protein expression in less accessible organs such as the placenta and uterus in this context.

There is an agreement on the fact that H_2_S is involved in PE, due to its physiological role in the regulation of blood pressure, angiogenesis, inflammation and oxidative stress [21,23]. However, literature reports on the exact pathophysiological role of H_2_S during PE remain inconclusive. Possomato-Vieira et al. report elevated plasma sulfide levels in PE patients compared to heathy pregnant women (N=120 (control) and 62 (PE), reported medians approx. 15-18 µM) [22]. In contrast, Wang et al. report reduced plasma sulfide levels in PE patients compared to heathy pregnant women (N=14 per group, reported medians approx. around 6-7 µM) [21]. Chaudhuri et al. also report reduced plasma sulfide levels in PE patients compared to healthy pregnant woment (N=100 per group, reported means 32.31 µM compared to 114.5 µM) [34]. All these studies used the methylene blue assay for sulfide detection. In contrast, in our study, both blood and plasma sulfide levels did not differ between case and control groups (medians around 0.1 µM) (see Figure 2). The methylene blue assay used in the abovementioned previous studies is generally considered to bear the risk of overestimating true sulfide levels, since numbers in the micromolar range are unlikely in biological samples [25,35]. Those levels are within in a potentially toxic range for the mitochondria [25]. Unfortunately, to the best of our knowledge, to date there is no “gold standard” for sulfide measurements in biological samples. We used a GC/MS based approach developed in house, which yields plasma sulfide values in the high nanomolar range. Furthermore, it is worthy of note that in both previous publications, there was a high variability in the measured values (range 3 – 35 µM [21], <1 – 100 µM [22], 32.31 ± 12.62 in PE patients and 114.50 ± 20.35 in controls [34]). For the first two studies this begs the question of the biological significance of the statistically significant median differences within a range of 1 – 3 µM [21,22]. The levels reported by Chaudhuri et al. are even much higher than the levels reported by Wang et al. and Possomoto et al., so that the data are difficult to compare with the latter. Taken together, we could not confirm any major difference in plasma sulfide levels between patients with PE and healthy pregnancies.

Elevated homocysteine levels (see Figure 2), in particular in ePE, still hint towards a dysregulation of endogenous H_2_S enzymes, in particular CBS, which uses homocysteine as a substrate. Our findings concerning homocysteine are in accordance with the literature: others reported a similar range of homocysteine levels and an elevation in the maternal serum of patients with severe pre-eclampsia compared to both mild pre-eclampsia and healthy pregnancies [36]. In both our and the cited study, levels of folate, Vit. B6 and Vit. B12, the lack of which can impair homocysteine catabolism, did not differ between groups (see Table 1, [36]), suggesting that other mechanisms are responsible for elevated homocysteine concentrations in the context of PE. Elevated homocysteine levels in early pregnancy have been reported to be able to serve as a predictor for the development of PE [37]. Furthermore, homocysteine levels were reported to decrease over the course of a healthy pregnancy [38]. This has been hypothesized to be related to increased metabolism by the placenta and other organs, which might be deficient in PE, thereby contributing to the elevated homocysteine levels seen in these patients [39]. Additionally, there are several SNPs in the CBS gene associated with PE [40]. We found elevated levels of CBS in both the placenta and the uterus of lPE, whereas there was no difference between ePE and control tissue CBS expression (see Figure 3 and 4). It is tempting to speculate that the up-regulated CBS expression in lPE normalized the homocysteine levels in this group, which did not happen in ePE, where CBS levels were unchanged and homocysteine was elevated.

Holwerda et al. also investigated placental expression of CBS and CSE in ePE and lPE. The authors found no differences in ePE or lPE with delivery matched controls for either CBS or CSE protein [23]. These findings are in contrast to our study, where we determined elevated CSE and CBS levels in lPE (Figure 3). However, we did not compare to delivery-matched controls: all our control and ePE patients had cesarean sections, whereas in the lPE group there was a rate of 39% vaginal deliveries (see Table 1). From Holwerda’s data it is obvious that a vaginal delivery was associated with lower placental CSE and CBS expression compared to cesarean section. Thus, the expectation for our study would be to see the lowest CSE and CBS levels in the lPE group, which, however, actually was the group with the highest levels (Figure 3), for CSE even confirmed with another method (Figure 5). Furthermore, we tested if there was a difference in placental CSE and CBS expression between spontaneous and cesarean deliveries in the lPE group, which was not the case (p=0.17 for CSE, p=0.80 for CBS, data not shown). Holwerda et al. do not clearly state which region of the placenta they focused on, whereas we focused on the intervillous space. Thus, the difference between their and our findings might be related to regional differences within the placenta. In contrast, Wang et al. determined down-regulated CSE protein in the placenta of PE patients, in accordance with the previously discussed reduced plasma sulfide levels they reported [21]. Here, the information on PE onset and delivery mode is unfortunately lacking, which makes it difficult to discuss their results in context with our data. Furthermore, the lower N=14 per group in their study [21] further limits the significance of their findings. Cindrova-Davies et al. confirm the findings of Holwerda et al. regarding the unchanged placental CBS expression in ePE compared to controls (all cesarean sections), but reported a different result for CSE: depending on the blood flow in the umbilical artery, CSE expression in the placental villi was either reduced (in PE with abnormal umbilical blood flow) or increased (in PE with normal umbilical blood flow) [24]. In our study, placental CSE expression was elevated in lPE, falling in with the previous data on PE patients with normal umbilical blood flow. However, in our study, umbilical blood flow was not documented, preventing a solid conclusion. Still, it is noteworthy, that various factors can impact placental expression of the endogenous H_2_S enzymes (PE, mode of delivery, umbilical blood flow), which should be considered in the design of future studies investigating the role of H_2_S (enzymes) during reproduction and parturition.

In the placenta, there was a direct linear relation of both CSE and CBS expression with OTR (see Figure 6), suggesting that there is an interaction of the H_2_S- and OT-system in the placenta and during PE. In the uterus, there was a direct linear correlation of CBS with OTR (see Figure 6). In sharp contrast, You et al. reported an inverse relationship of CSE and CBS with contraction-associated proteins, such as OTR, in the pregnant laboring myometrium: in myometrial samples from women undergoing cesarean section after going into labor, CSE and CBS were reduced while OTR was elevated compared to tissue samples of women that had not gone into labor before the cesarean section [17]. In our study, the control group only comprised patients with scheduled cesarean section, thus we did not investigate samples from patients in active labor. In addition, 39 % of patients in the lPE group had a vaginal delivery (see Table 1) and thus went into labor, however, for obvious reasons, no myometrial samples from these patients were available. Thus, it is difficult to compare You et al.’s results with our data. However, the inverse relationship of CSE/CBS with OTR reported by You et al. seemed to be strongly mediated by H_2_S, since administration of both NaHS and L-cysteine (as a substrate for both CBS and CSE) induced a significant dose-dependent reduction of OTR expression in uterine smooth muscle cells [17]. NaHS is known to be able to release high peak sulfide concentrations [25], which were most likely also present in the *ex vivo* study by You et al. In our study, we did not detect any difference in circulating sulfide levels between groups (see Figure 2) and endogenous sulfide levels are much lower than with NaHS administration, which might also explain why we observed a different relationship of CSE/CBS with OTR.

Surprisingly, despite the well-established function of OTR during parturition, data on its role in PE are very scarce. In contrast to our observation of elevated OTR in the placenta in lPE compared to controls (see Figure 3), Fan et al. reported a reduction of OTR mRNA and protein with PE [20]. However, the authors specifically sampled the main stem villous arteries for their analyses [20], whereas we quantified OTR expression in the intervillous space, which mostly contains the terminal villi. This might help explain the difference in results between that report and our data. Furthermore, placental OTR expression increases with gestational age [18], which might be a confounder in this context, since PE is normally associated with gestationally earlier deliveries: in both Fan et al.’s (control: 38.1 weeks, PE: 33.5 weeks) and our study (control: 38.8, ePE: 29.6 weeks, lPE 36.8 weeks), gestational age at delivery was significantly lower with PE compared to control (see Table 1, [20]). However, the difference in gestational ages between control and lPE in particular is less drastic than in the study by Fan et al., suggesting that the confounding effect of gestational age would be more significant in their than in our study. Furthermore, it has been suggested that OT/OTR signaling mediates myometrial contractility during parturition, since patients with cesarean section had lower placental OTR expression compared to placentas from spontaneous deliveries [41]. It is difficult to interpret our findings in context with this report: patients in our control and ePE groups all had cesarean sections, which seems to be associated with lower placental OTR *per se*. In the lPE group 39% of patients had a vaginal delivery, which would potentially be associated with higher average OTR expression: that is exactly what we observed in our study. However, when we compared placental OTR expression in just the lPE group separated by mode of delivery, no significant difference could be detected (p=0.18, data not shown). In contrast, in the study by Fan et al., all patients had a vaginal delivery [20], suggesting that the placental OTR expression they detected was higher overall compared to the patients in our study. Unfortunately, we did not collect placentas from healthy pregnancies with vaginal deliveries, thus we cannot really correct our data for mode of delivery. All in all, considering that mode of delivery and gestational age both may represent potential confounders for placental OTR expression, no clear conclusions on its role during PE can be drawn from the data presented here and in the literature.

NO is a downstream signaling mediator of the interaction of H_2_S and OT [15,42]. In the literature, both a direct [34] and inverse [22] relation between plasma sulfide levels and nitrite/nitrate as correlates of NO availability have been reported in PE patients. Interestingly, in one study, the direct correlation of sulfide with NO availability was also present in the control group, i.e., healthy pregnant women [34], whereas in the study that found the inverse relation in PE, there was no correlation in healthy pregnant women [22]. The contrasting findings in the two studies might be related to the different methodology used to determine NO availability (Griess reaction [34] *vs.* NO-Analyzer [22]). Interestingly, both studies agree on the fact, that PE is associated with lower overall levels of NO in PE patients, which has also been reported in the literature previously [43]. Furthermore, PE is reportedly associated with reduced plasma and placental levels of the constitutive NO-releasing enzyme endothelial NO-synthase (eNOS) [44,45]. We did not determine eNOS expression in samples from our patients. However, we evaluated nitrotyrosine formation, which critically depends on NO-availability and simultaneous presence of superoxide [26]. Moreover, pregnancy and parturition are both associated with the occurrence of oxidative stress [46]. Patients with PE are characterized by elevated markers of oxidative stress and reduced antioxidant enzyme activities compared to healthy pregnant women [47]; in addition, oxidative stress is considered to contribute to endothelial dysfunction in the placenta [48]. Thus, it is not surprising that we observed nitrotyrosine formation as a tissue marker of oxidative stress in both the placenta and uterus of all patients and most pronounced in PE (see Figure 3 and 4). In line with these results, we observed an induction of HO-1 expression in PBMC of PE patients (see Figure 5), inasmuch as it could represent a compensatory antioxidant mechanism. HO-1 expression can be stimulated *via* the transcription factor Nuclear response factor 2 (Nrf2) [49], which in turn can be activated by H_2_S signaling [50] and might thus represent a point of interaction of these systems, as has been previously reported in basic research in atherosclerosis [51]. Furthermore, NO can induce HO-1 expression during stress conditions [52–54], which is another potentially relevant molecular mechanism in the pathophysiology of PE. So far, no differences in serum HO-1 levels have been reported between control and PE patients [55]. However, the same study also reported significantly lower plasma HO-1 in mild PE compared to severe PE [55]. A low N and high variability of HO-1 expression in PBMC of ePE patients make it difficult to interpret our results for this group compared to lPE. Moreover, it is noteworthy that in the study by Sandrim et al., blood was sampled at 20-25 weeks of pregnancy [55], whereas in our study, blood for PBMC isolation was sampled at the timepoint of recruitment, which was associated with a higher mean gestational age (35 weeks + 3 days; 28 weeks + 3 days for ePE, significantly earlier than in the other two groups; 35 weeks + 5 days for lPE, 36 weeks + 2 days for control), potentially confounding our results for PBMC protein expression. Literature data on the effect of gestational age on HO-1 expression are in fact controversial: in the placenta, both no effect [56] and an increased expression [4] of HO-1 with gestational age have been reported. Reports on the role of placental HO-1 during PE are equally controversial [57]: either a reduction of placental HO-1 in PE [4], or no difference in placental HO-1 expression in PE compared to controls [58–60], or higher expression with unchanged activity [61] have been reported. Our results are well in line with the literature reports of unaffected placental HO-1 expression in PE (see Figure 5), which also might question the hypothesis that HO-1 assumes major importance as an antioxidant response during PE.

## Limitations

The study presented here is limited by the fact that the N of recruited patients in the ePE case group was rather low, since separating the case group into the ePE and lPE subgroups was only performed post hoc and not part of the initial study design. Furthermore, we only investigated the placental intervillous space, since the chorionic and basal plate were not consistently available from the sampled specimens. Last but not least, we had an unexpectedly high number of vaginal deliveries within the lPE group. This might confound the results, especially when comparing to the other study groups, in which all patients had a cesarean section. We addressed this issue by testing if there were significant differences in placental protein expression between patients with cesarean section and vaginal delivery in the lPE group, which was not the case (data not shown).

## Conclusion

In this observational case control study, there were significantly elevated systemic homocysteine levels and significantly elevated levels of endogenous H_2_S enzymes and OTR in the placenta of patients with pre-eclampsia, in spite of unchanged systemic sulfide levels. These findings suggest a role for the interaction of the endogenous H_2_S- and OT/OTR systems in the pathophysiology of pre-eclampsia, possibly linked to impaired antioxidant protection.

## Data Availability

All data produced in the present study are available upon reasonable request to the authors.

## Author Contributions

Conceptualization, P.R., and C.W.; methodology, T.M., O.M., U.W., and T.B.; validation, T.M., O.M., U.W., and T.B.; formal analysis, T.M. and O.M.; investigation, S.E., N.D., S.K., U.W., C.F., C.B., J.T., and T.B.; resources T.P., C.B., P.R., and C.W.; data curation, T.M., and E.R.; writing—original draft preparation, T.M.; writing—review and editing, P.R., and C.W.; visualization, T.M.; supervision, O.M., C.W., T.B., P.R., and C.W.; project administration, P.R. and C.W.; funding acquisition, T.M. All authors have read and agreed to the published ver-sion of the manuscript.

## Institutional Review Board Statement

The study was conducted in accordance with the Declaration of Helsinki, and approved by the Landesärztekammer Bayern (file number 19033, 29^th^ August 2019).

## Informed Consent Statement

Informed consent was obtained from all subjects involved in the study.

## Data Availability Statement

The raw data supporting the conclusions of this article will be made available by the authors on request.

## Acknowledgments

We would like to thank Rosy Engelhardt, Andrea Seifritz, Rosie Mayer, Bettina Stahl and Vittoria Passarelli for their skillful technical assistance.

